# Imageless navigation system (Naviswiss) provides accurate component position in total hip arthroplasty with lateral decubitus position for end-stage hip osteoarthritis: A prospective cohort study with CT-validation

**DOI:** 10.1101/2023.06.05.23289691

**Authors:** Corey Scholes, Manaal Fatima, Tobias Schwagli, David Liu

## Abstract

**Aims:** The Naviswiss system (Naviswiss AG, Brugg, Switzerland) is a handheld imageless navigation device used to improve the accuracy of implant positioning in total hip arthroplasty (THA). However, clinical data for leg length discrepancy and femoral offset is lacking, and the validity of the system has not been reported for patients undergoing THA in the lateral decubitus position. This study aimed to report the accuracy of the device in this patient population.

**Methods:** Patients underwent THA in the lateral decubitus position by a single surgeon. Component positioning measured by the device intraoperatively was compared to postoperative measurements on computed tomography (CT) scans. Agreement between the navigation system and postoperative measurements was reported with respect to acetabular cup inclination, acetabular cup version, femoral offset and leg length discrepancy.

**Results:** The mean difference between intraoperative and postoperative CT measurements was within 2° for angular measurements and 2mm for leg length. Absolute differences for the two indices were within 5° and 4mm. Mean bias was 1-2° overestimation for cup orientation and up to 2mm overestimation for leg length change, but 95% limits of agreement did not exceed absolute thresholds of 10° and 10mm, especially after correction for bias. Four cases (12%) were declared intraoperatively for issues with fixation on the greater trochanter. While inclusion of these cases generated acceptable accuracy overall, their omission improved between-case variability in accuracy and reduced the LOA for both offset and leg length.

**Conclusions:** The accuracy of the Naviswiss system falls within clinically acceptable recommendations for acetabular cup placement, femoral offset and length length. The system could be further improved with regression-based bias correction.

## Introduction

Accurate component placement is an important factor affecting outcomes of total hip arthroplasty (THA) (Davenport & Kavarthapu, 2016). Inappropriate positioning of the femoral stem or acetabular cup is associated with impingement, instability, bearing failure, leg length discrepancy and an increased risk of the need for revision surgery (Domb et al., 2015; Liebs et al., 2014; Renkawitz et al., 2016).

Efforts to guide component placement in THA include robotic-assisted surgery and computer navigated systems (Chen et al., 2018). Computed tomography (CT)-based navigation relies on preoperative planning on CT images which increases both cost and radiation exposure, while imageless navigation relies on landmark registration and does not take into account detailed patient-specific anatomy (Sugano, 2013). Studies have shown imageless navigation to increase the accuracy of acetabular implant position (Migliorini et al., 2022), but surgical time remains markedly longer (Kunze et al., 2022) with no clinically meaningful differences in patient-reported outcomes (Singh et al., 2021). Despite being in use for over 20 years, the expenses associated with the purchasing and maintenance of navigation systems as well as the usability and longer operation time have prevented their widespread use (Renner et al., 2017).

Advances in imageless technology have seen the development of portable navigation systems including accelerometer-based devices, which have been advantageous in improving the accuracy of implant positioning (Shigemura et al., 2021) and tested with various surgical approaches (Bradley et al., 2019; Grosso et al., 2016; Hayashi et al., 2019). The Naviswiss system (Naviswiss AG, Brugg, Switzerland) is a handheld imageless navigation device that utilises an infrared stereo camera and an inertial measurement unit to calculate the position and orientation of the pelvis, greater trochanter and camera in space (Ektas, Scholes, Ruiz, et al., 2020). It has been tested in vivo with an anterolateral approach in the supine position, with <3° mean absolute error for cup inclination and anteversion (Hasegawa et al., 2022). The system has also been tested using a direct anterior surgical approach with fluoroscopy, and the absolute difference between intraoperative and radiographic measures was <3.5° for cup orientation (Ong et al., 2022). However, clinical data for leg length discrepancy and femoral offset is lacking, and the accuracy of the system has yet to be reported for patients in the lateral decubitus position. Hip navigation systems have in the past have been difficult to use with hip approaches in the lateral decubitus posture due to issues with registering the anterior pelvic plane.

A trial protocol was developed to assess the validity of this new and unique hip navigation system in measuring acetabular cup inclination, acetabular cup version, femoral offset and leg length discrepancy (Ektas, Scholes, Harrison-Brown, et al., 2020). This study reports the accuracy of the device by comparing intraoperative component positioning to postoperative CT measurements, specifically in patients undergoing THA using an anterolateral approach in the lateral decubitus position. We hypothesised the handheld navigation system would provide precise intra-operative information for acetabular cup orientation, as well as leg length and offset change following primary THA in the lateral decubitus position.

## Methods

### Patient selection

Ethical approval for the study was obtained from the Ramsay Health Care Queensland Human Research Ethics Committee (HREC Reference 20/08), and the study was registered with the Australian New Zealand Clinical Trials Registry (ACTRN12620000873921). Patients over 18 years of age were invited to participate in the study if they presented to the participating surgeon with end-stage osteoarthritis or inflammatory arthritis, and underwent THA surgery with an anterolateral surgical approach in the lateral decubitus position.

Patients were excluded from the study if they: were unable to provide informed consent; had declined or revoked consent for use of clinical data for the study; had severe contralateral hip deformity or dysplasia; required a simultaneous bilateral procedure; required an ipsilateral revision procedure; had a short-stem component implanted; or were lost to follow-up.

### Surgical technique and intraoperative measurement of component positioning

All patients underwent THA by a single surgeon using identical surgical technique and perioperative protocol. Preoperative templating was performed using 3D CT assessment and an automated 3D planner (Formus Labs, Auckland, New Zealand). The 3D planner predicted the femoral component according to endosteal fit within the femur and aimed for combined anteversion of 37° by adjusting the cup version.

Patients either received a spinal anaesthetic with sedation or general anaesthetic, or general anaesthetic alone with muscle relaxant under the discretion of the anaesthetist. The patients were positioned in the lateral decubitus position and held stable with standard hip bolster supports at the sacrum and anterior superior iliac spines. The pelvic position was checked by the operating surgeon to ensure a stable vertically parallel anterior pelvic plane prior to draping. Two 3mm pins were inserted into the iliac crest to secure the pelvic tracker and a mini-plate secured with a single 3.5mm locking screw of 25mm in length was used to attach the femoral tracker to the greater trochanter of the femur. The femoral tracker could be removed and reattached to the locking plate when needed for leg length and offset measurement. The Naviswiss camera was draped and secured to a small trolley on the contralateral side to the surgeon. The camera was operated by the surgical assistant.

The hip was exposed through an anterolateral approach detaching the anterior 50% of the abductor insertion just anterior to the musculotendinous junction, keeping the gluteus medius and minimus as one flap. All landmarks were registered according to the standard workflow including the hip centre of rotation (COR), anterior and posterior attachments of the transverse acetabular ligament and superior acetabular rim. The COR is identified by the functional method (Leardini et al., 1999), with the thigh moved through a multiplanar range of motion while the tags are tracked with the handheld camera. The hip was dislocated anteriorly and following bone preparation, uncemented acetabular and femoral components were inserted. All patients received an E1 Poly liner and ceramic head. Cup position was registered after stable impaction on the acetabular shell. Final length leg and offset was measured after the hip was relocated with the real femoral head. The final intra-operative component positions were logged by the navigation system and exported for analysis: Acetabular cup inclination (ACI), acetabular cup version (ACV), femoral offset (FO) and leg length discrepancy (LLD). All surgeries were performed by the senior author.

### Measurement of component positioning

The primary study outcomes were extracted for analysis as previously described (Ektas, Scholes, Ruiz, et al., 2020). Agreement between the navigation system applied intraoperatively and postoperative measurements was assessed using the following parameters:

- ACI - The angle between the acetabular and longitudinal axes when projected onto the functional pelvic plane (FPP)
- ACV - The angle between the acetabular axis and the FPP
- FO - The relative difference between the hip centre of rotation of the operated joint relative to its starting position at the initial assessment in the coronal plane (medial-lateral) within the pelvic coordinate system
- LLD - The change in the distance between the greater trochanter tag and the hip centre of rotation summed with the change in the distance between the centre of the acetabulum and the centre of the cup in the transverse plane (superior-inferior)

Blinded images in DICOM (Digital Imaging and Communications in Medicine) format were used for all pre- and postoperative CT measurements of component positioning, with information relating to the specific diagnosis, study, surgeon or whether navigation used for the hip arthroplasty procedure removed prior to measurement of component position. The pre- and postoperative images were blinded by an independent research assistant who was not involved in performing the measurements.

Postoperative component position was measured by loading the DICOM data to dedicated software (3D Slicer, www.slicer.org) to measure version and inclination of the acetabular cup. FO and LLD were measured through assessment of anatomical landmarks picked in pre- and postoperative scans. For the postoperative CT assessments, coordinate systems for the pelvis and femur were determined based on the anatomic landmarks. Parameters from both the Naviswiss and CT analysis system were expressed relative to the FPP, with the origin placed at the centre of the line connecting the left and right ASIS. For the postoperative CT analysis, the position of the cup centre was compared with the native hip COR determined from the preoperative CT. FO and LLD was reported as the pre-to-post change of the femoral coordinate frame relative to pelvis FPP coordinates in the coronal (mediolateral) and transverse (inferior-superior) planes respectively.

## Data and Statistical Analysis

### Missing data

One case was missing height and weight - replacement with group mean for body mass index (BMI) was used to address the missing value to enable regression on the full dataset.

### *Summary agreement* (uncorrected and declarations included)

Mean deviation (delta) was calculated by subtracting the imaging measurement from the intraoperative measurement. A positive result indicated an overestimation by the navigation system, and a negative value denoted an underestimation. A bootstrap analysis for delta was performed for each measurement (ACI, ACV, FO, LLD) with 1000 replications with replacement and a fixed initial seed. Mean, standard error, standard deviation, 95% confidence intervals for the mean were calculated. A Bland-Altman plot was generated for each measure using the with limits of agreement (LOA) calculated with the formula (Bland et al., 1999; Giavarina, 2015);

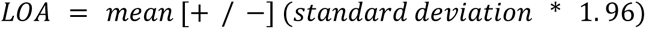

Linear regression was used to assess the relationship between mean deviation (delta) and average of the intraoperative and imaging measurements.

Delta was converted to absolute values and the summary statistics calculated with the bootstrap analysis as described.

### Bias assessment and correction

Linear regression with bootstrapping was used to assess the relationship between delta and the intraoperative measurement, adjusted for age at surgery, BMI and sex. Model predictions based on the original intraoperative measurements were used to create a bias-adjusted version of the intraoperative measurements and a bias-corrected delta calculated by subtracting the imaging measurements from the bias-corrected intraoperative measurements. Alternative correction for offset and leg length change was performed by dropping values where an intraoperative declaration was made and the summary agreement analysis repeated as described above (Supplementary material 1). All statistical analyses were performed using Stata (v17.1, Statacorp, USA), with alpha set at 5% to indicate significant effects where appropriate.

## Results

### Patient characteristics

A cohort of 109 consecutive cases undergoing primary THA were assessed for eligibility with 33 included for study analysis (Figure 1). The analysis cohort comprised 52% females and had a mean age at surgery of 68.9 years (SD 8.8) and a mean BMI of 28.2 (SD 4.7).

**Figure 1:**
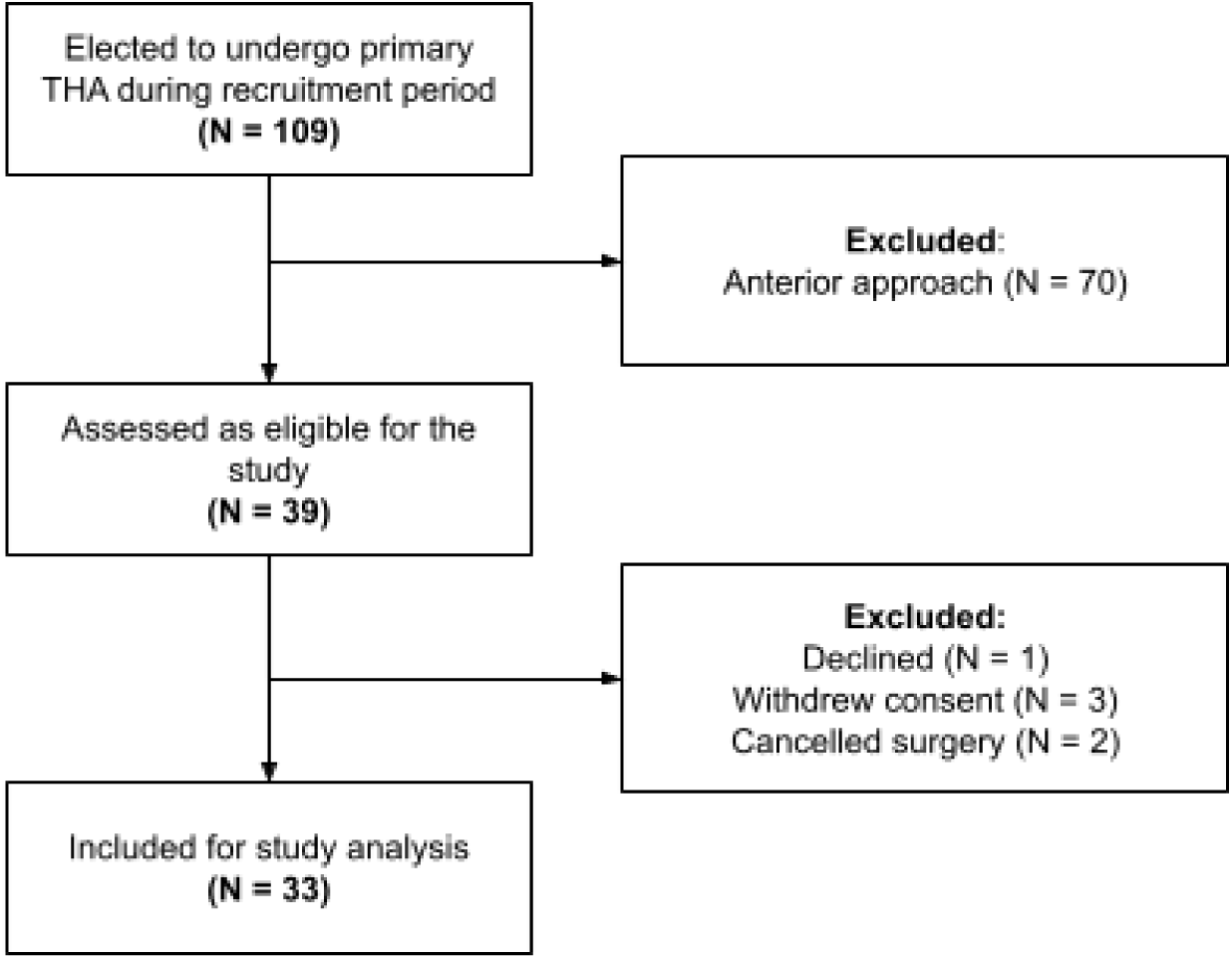
STROBE diagram (von Elm et al., 2007) of patient inclusion into the study analysis

### Complications

Complications were observed in four cases - a periprosthetic infection that underwent washout with liner and head exchange at 8 weeks; stem subsidence that stabilised after 5 months and did not require further surgery and was symptom free; a superficial wound infection that resolved with a course of oral antibiotics and one case of numbness in the contralateral thigh that resolved without intervention.

### Agreement - Intraoperative to Imaging

#### Thresholds and declared observations

Intraoperative declarations were made for six patients, with loss of fixation of the tracker on the greater trochanter (n = 4) and acquisition failure (n = 2) the reasons cited. One patient without an intraoperative declaration exceeded the specified measurement thresholds for both inclination and version, but not for offset or LLD (Table 1).

**Table 1:**
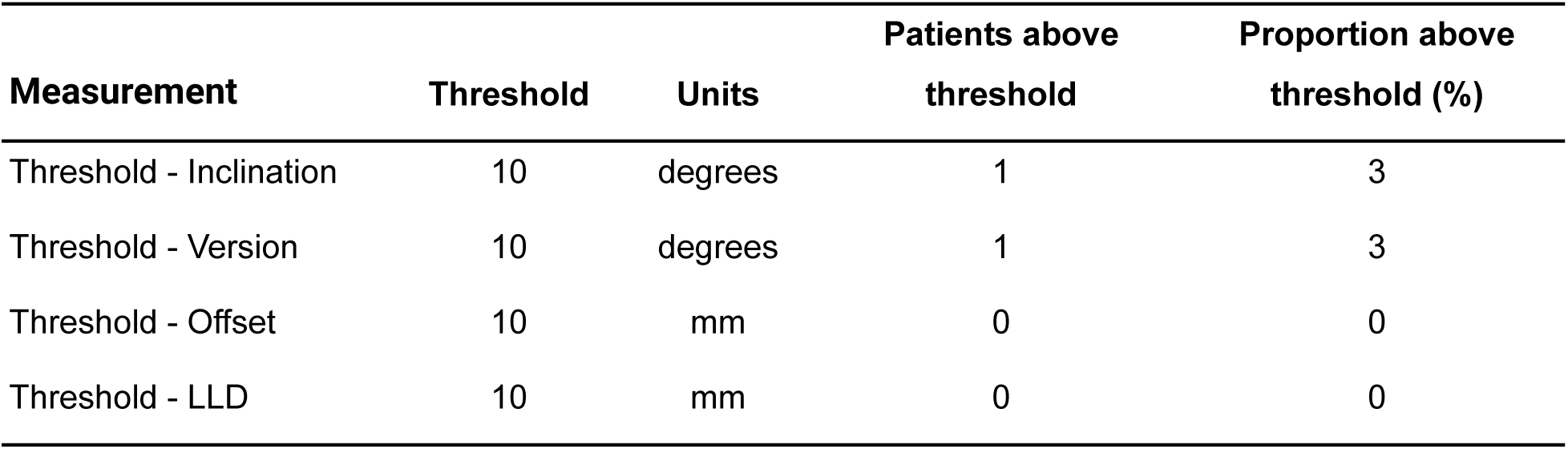
Patients without an intraoperative declaration exceeding the measurement thresholds

The mean differences between the intraoperative measurements and the postoperative imaging analysis were less than one degree for inclination, less than two degrees for version, and less than 2 mm for both offset and LLD (Table 2). The 95% LOA were within 10 degrees for inclination and version, and within 8 and 6mm for offset and LLD respectively (Table 2, Figure 2). Overall, 90% of cases (95%CI 75.3 - 98.1) were within 10° of the image-measured for both inclination and version (Figure 3). The linear fit of the average to the delta indicated that the bias between the navigation and the CT measurements was not constant across magnitude for inclination, version and LLD (p<0.001, Table 3).

**Figure 2:**
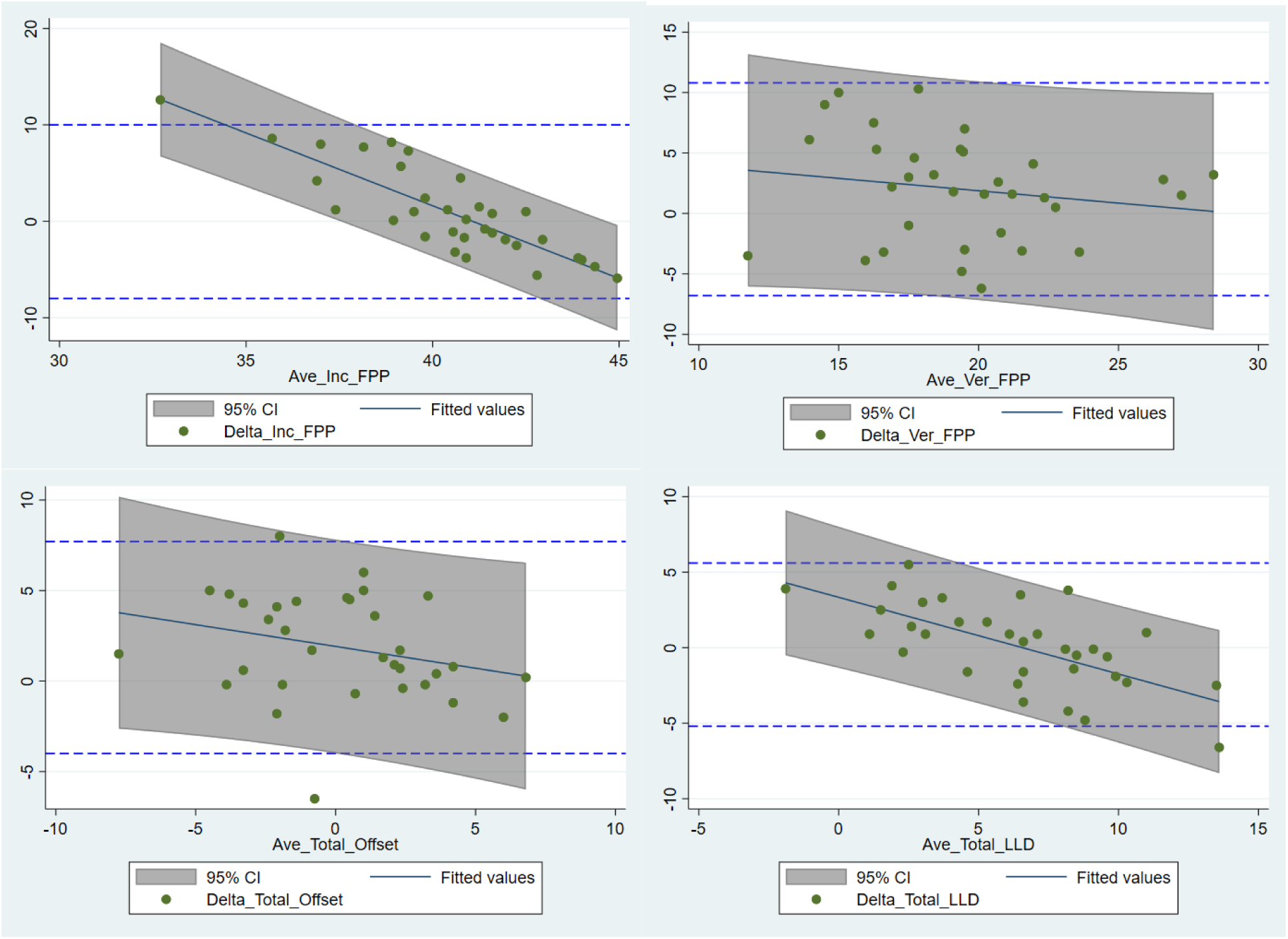
Bland-Altman plots with 95% limits of agreement for inclination and version, offset and leg length (LLD). Regression fits with shaded areas denoting 95% prediction intervals indicate the relationship between magnitude and agreement.

**Figure 3:**
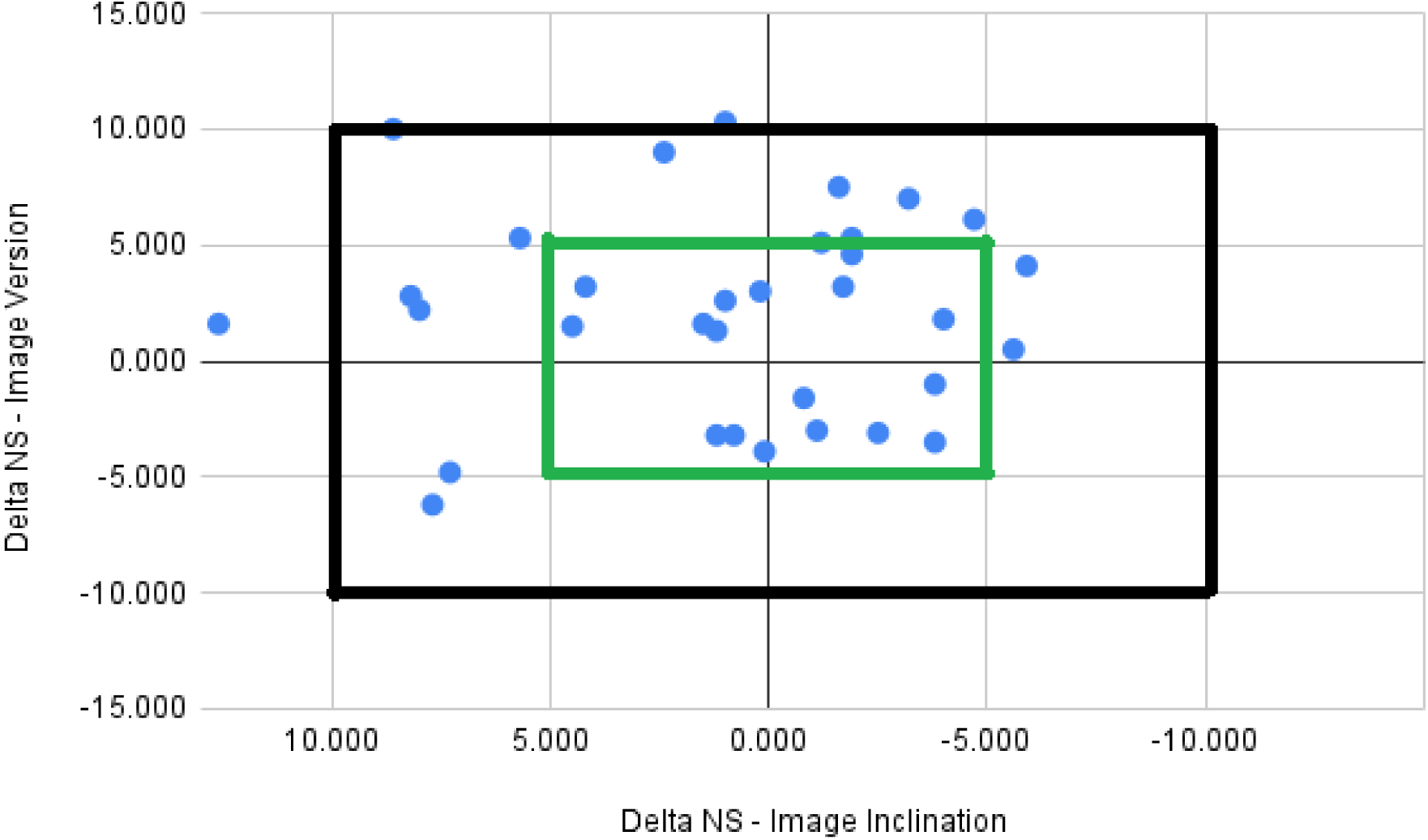
Scatterplot of delta in version versus inclination for all cases. Outer box - 10° threshold; Inner box - 5° threshold.

**Table 2:**
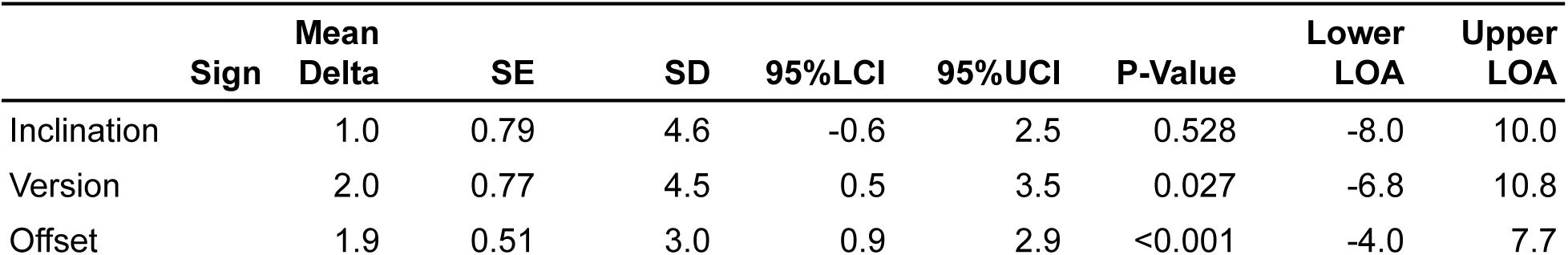
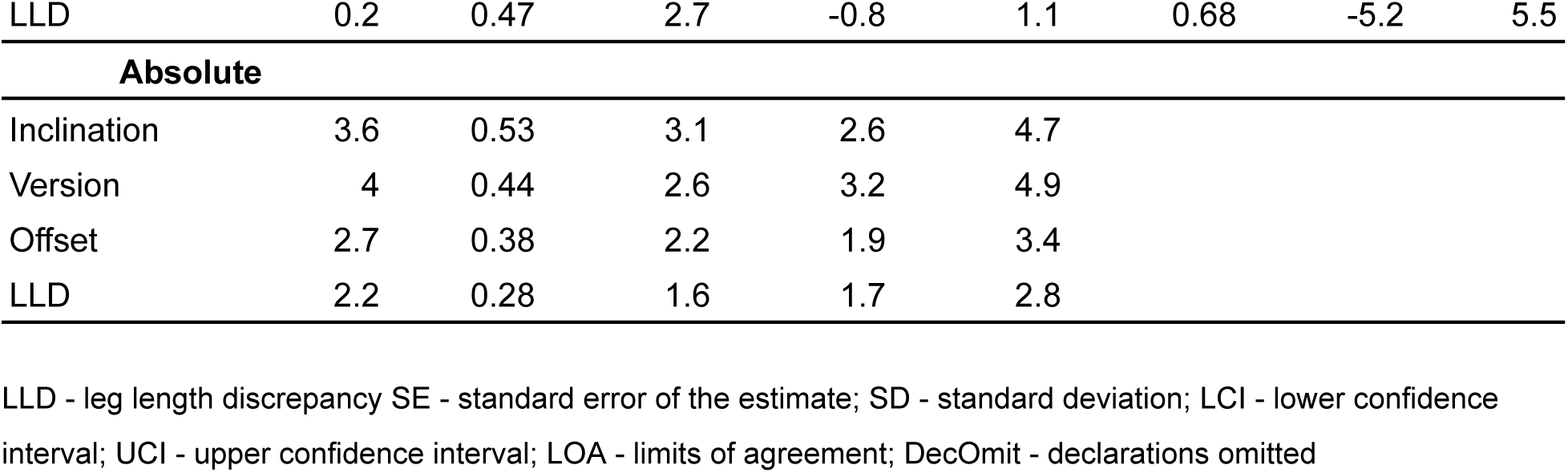
Summary of mean differences between intraoperative and image-based measurements. P-value indicates probability of observing mean delta (relative to zero) as extreme assuming the null hypothesis is true

**Table 3:** Linear fit of average of measurements to delta of measurements

|  | **Coefficient** | **SE** | **95%LCI** | **95%UCI** | **P-Value** | **Adjusted R-sq** |
| --- | --- | --- | --- | --- | --- | --- |
| Inclination | -1.51 | 0.1 | -1.70 | -1.31 | <0.001 | 0.70 |
| Version | -0.2 | 0.23 | -0.65 | 0.24 | 0.368 | 0.00 |
| Offset | -0.24 | 0.15 | -0.53 | 0.05 | 0.102 | 0.04 |
| LLD | -0.51 | 0.09 | -0.69 | -0.32 | <0.001 | 0.41 |

### Factors associated with agreement and bias correction

BMI, age and sex were tested as main effects against all measurements of interest. The regression results indicated a significant association (P = 0.049) of sex with inclination delta, a magnitude-dependent bias for version (P = 0.005) and offset delta (P = 0.011) (Supplementary material 2). Bias correction applied to the intraoperative measures removed overall bias and shrank the between-case variation (SD) of delta by 6-16%, which increased to 7-42% for absolute values (Table 4). Bias correction also shrank the mean absolute delta by 5-26% relative to the uncorrected values. Alternatively, by omitting declared observations for offset and leg length (Table 5), mean absolute error was reduced by 18-33% and between case variability by 18-39%.

**Table 4:** Summary of mean differences between intraoperative and image-based measurements for bias corrected intraoperative measures

| **Sign** | **Mean Delta** | **SE** | **SD** | **95%LCI** | **95%UCI** | **Lower LOA** | **Upper LOA** |
| --- | --- | --- | --- | --- | --- | --- | --- |
| Inclination | 0 | 0.69 | 4.0 | -1.4 | 1.4 | -7.9 | 7.9 |
| Version | 0 | 0.69 | 4.0 | -1.4 | 1.4 | -7.9 | 7.9 |
| Offset | 0 | 0.43 | 2.5 | -0.8 | 0.8 | -4.9 | 4.9 |
| LLD | 0 | 0.44 | 2.6 | -0.9 | 0.9 | -5.0 | 5.0 |
| **Absolute** |  |  |  |  |  |  |  |
| Inclination | 3.2 | 0.47 | 2.7 | 2.24 | 4.08 |  |  |
| Version | 3.3 | 0.37 | 2.2 | 2.6 | 4 |  |  |
| Offset | 2 | 0.22 | 1.3 | 1.58 | 2.45 |  |  |
| LLD | 2.1 | 0.26 | 1.5 | 1.58 | 2.59 |  |  |

**Table 5:**
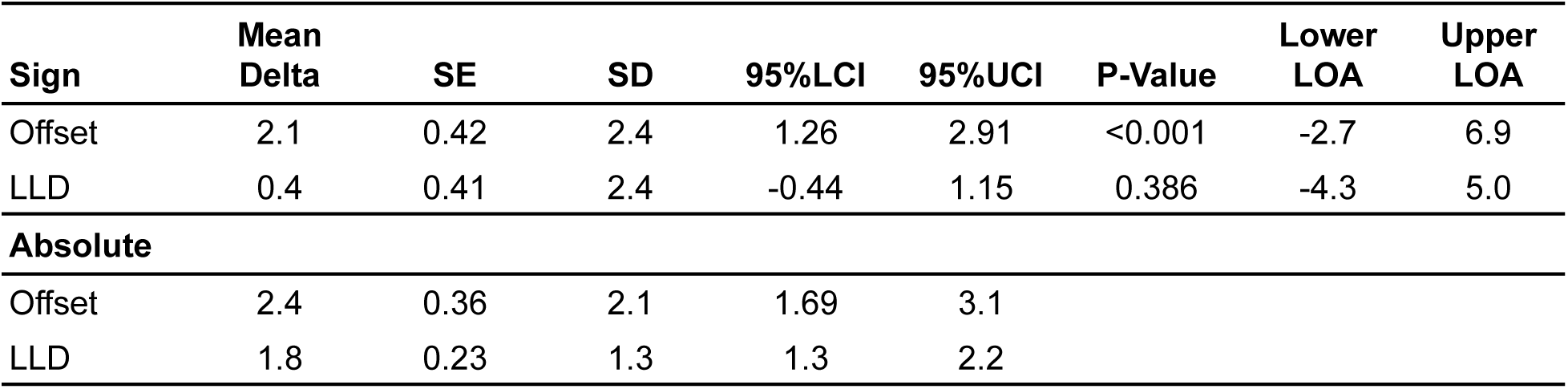
Summary of mean differences between intraoperative and image-based measurements with declaration cases omitted (N = 27)

| **Sign** | **Mean Delta** | **SE** | **SD** | **95%LCI** | **95%UCI** | **P-Value** | **Lower LOA** | **Upper LOA** |
| --- | --- | --- | --- | --- | --- | --- | --- | --- |
| Offset | 2.1 | 0.42 | 2.4 | 1.26 | 2.91 | <0.001 | -2.7 | 6.9 |
| LLD | 0.4 | 0.41 | 2.4 | -0.44 | 1.15 | 0.386 | -4.3 | 5.0 |
| **Absolute** |  |  |  |  |  |  |  |  |
| Offset | 2.4 | 0.36 | 2.1 | 1.69 | 3.1 |  |  |  |
| LLD | 1.8 | 0.23 | 1.3 | 1.3 | 2.2 |  |  |  |

## Discussion

The main finding of our study is that the handheld hip navigation system (Naviswiss) provides information that can assist accurate acetabular cup orientation and leg length and offset restoration in the lateral decubitus position. The mean difference between intraoperative and postoperative CT measurements was within 2° for angular measurements and 2mm for the length measurements. The absolute differences for the two indices were within 5° and 4mm.

The mean absolute deviation of acetabular inclination (3.6, 95%CI 2.7 - 4.5) between the navigation system and the CT-based analysis was comparable to the deviation reported by (Naito et al., 2022) (2.8, 2.3 - 3.3) with patients in the supine position, but higher than the pooled deviation (2.6, 2.4 - 2.8) for previous studies in the lateral decubitus patient position using CT-based, imageless and accelerometry systems (Supplementary material 3 - summary of validation findings). In contrast, anteversion mean absolute deviation (4, 3.1 - 4.9) was greater than Hasegawa (2.8, 2.3 - 3.3), but comparable to pooled deviation for previous studies (3.6, 3.4 - 3.8). Overall, mean bias was 1-2° overestimation for cup orientation and up to 2mm overestimation for leg length change, with 95% LOA on or below 10° for orientation and 5-10mm for offset/leg length change. Absolute thresholds of 10° and 10mm were established a-priori in the study protocol (Ektas, Scholes, Ruiz, et al., 2020) and were not exceeded by 95% LOA, especially after correction for bias. Between-patient variation in published guidelines for cup orientation vary between 5 and 12° for inclination and up to 18° for version (Harrison et al., 2014). In general, less than 10mm of LLD is considered acceptable after THA (McWilliams et al., 2013). In addition, a simulation study (Shoji et al., 2018) reported impingement and loss of motion range with a 4mm medialisation/lateralisation of the cup, although this amount of change was not justified in their methods. The LOA in the present study suggest that the entire patient sample would fall within these tolerances, except for offset, where the 95% LOA exceeded 4mm. In reality the clinical tolerance for error will not be distributed evenly across the population, with patients at the extremes (smaller or larger) of target orientation and position requiring greater accuracy to prevent more extreme cup positions.

Comparing the present results to the literature should be done with caution, due to the heterogeneity of case-mix, the systems employed for intraoperative measurements, the methods by which gold-standard data was derived (CT vs radiograph), as well as analytical methods (pelvic coordinate systems, statistical analysis) and reporting standards. Recent meta-analyses have reported substantial differences in patient demographics, surgical technique (Migliorini et al., 2022) and outcome heterogeneity (Shigemura et al., 2021) in studies comparing navigated THA to conventional instrumentation. In addition, some validation attempts have used the anterior pelvic plane (APP) coordinate system (Hasegawa et al., 2020; Yamada et al., 2018), compared to the FPP used in the present analysis. Further, due to the proprietary nature of the systems under consideration, the presence or absence of bias-correction or other real-time compensatory calculations in the system software is not reported in all studies. Nevertheless, the reasons for higher deviation reported in the present study compared to other validation reports in the literature may be related to differences in case-mix between studies. Foremost are differences in BMI and indication for surgery. (Hasegawa et al., 2022) reported on accuracy in a cohort of patients with an average BMI of 24.6±4.9 compared to the present study 28.2±4.7. Other sources of error include the validity and reliability of the CT measurements which have up to 2-3° of deviation from true (Schwarzkopf et al., 2017), despite high relative reliability (Hasegawa et al 2021). In addition, the intraoperative measurements from the system are rounded to the nearest whole number, which could create up to 1° deviation from the image-based measurement.

A biphasic pattern of magnitude-dependent bias (Ho, 2018) was observed for inclination (FPP) and leg length change. The navigation system tended to overestimate at smaller average measurements and underestimate at larger averages (Figure 2). While bias correction was able to re-centre the sample around zero and reduce between-patient variation, further work is needed to validate regression-based bias correction algorithms to remove the magnitude-dependency (slope) by adjusting intraoperative measurements, potentially based on patient demographics and/or other factors. In addition, further work may be required to improve tracker fixation for offset and leg length with 4 cases (12%) declared intraoperatively for issues with fixation on the greater trochanter. While inclusion of these cases generated acceptable accuracy overall, their omission improved between-case variability in accuracy and reduced the LOA for both offset and leg length, highlighting a potential avenue for expanding patient indications. A previous study (Hasegawa et al., 2022), mentioned the potential vulnerability of the system to pin fixation on the iliac crest.

The magnitude-dependent bias observed for inclination in the present series may be associated with higher BMI compared to previous studies. The soft-tissue distribution over key bony landmarks of the pelvis, with particular reference to the shift in distribution with changes in patient position during measurement and surgical approach may be a key source of error in this context. Primarily, a thicker layer of soft tissue at key landmarks may dampen intraoperative measurements and limit the sensitivity of the system to pelvic anatomical variation. A similar study conducting a CT-validation of an accelerometer-based navigation system reported a significant correlation between cup orientation delta and BMI (Hayashi et al., 2019), in a sample with lower average BMI (23.7±4.4). Soft-tissue distribution is also related to age and sex (He et al., 2018), and may contribute to the systematic overestimation of inclination for females in the present series.

A limitation of navigation in hip replacement surgery is dependent on landmark acquisition, which is itself dependent on patient BMI, soft-tissue and surgical setup (including draping). The system used in the current study is reliant on stable accurate pelvic positioning, which can be a challenge for the surgeon to replicate the FPP established on the CT 3D-reconstruction by visualising the bony landmarks on the operating table. The ability to identify landmarks and the assumptions made about the relationship between pelvic orientation and those landmarks is key to aligning the plane on the table to that generated by the image-based analysis.. Like many previous navigation systems, the Naviswiss relies on the FPP for version and inclination values. However the FPP can be difficult to measure in the lateral decubitus position and alignment between the measured plane (detected by the trackers) and the true plane may diverge during the various steps of the THA procedure during retraction and leg positioning for exposure. Others have alluded to the effect of soft tissue thickness over key pelvic landmarks (Lee & Yoon, 2008; Ybinger et al., 2007), albeit mitigated by surgeon experience (Ybinger et al., 2007). Still others have observed challenges of landmark identification after draping (Lin et al., 2010), and this combined with greater tissue thickness may have reduced the sensitivity of the system. Soft-tissue distribution is also related to age and sex (He et al., 2018), and may contribute to the systematic overestimation of inclination for females in the present series. Interestingly this was not the case in the Hasegawa series - where 90% of the cohort were female. The reasons for this effect in the present study is not clear, however correction for this bias in the intraoperative measurements reduced mean absolute deviation.

The present study describes CT-based validation of a navigation system for THA in end-stage osteoarthritis, with *horizontal* LOA between intraoperative and CT measurements of inclination, anteversion, offset and leg length. However, the results should be placed in context of key limitations inherent in the study. The first is that the results indicate magnitude-dependent bias within the tolerance thresholds set a-priori, undermining the efficacy of summary statistics for delta and horizontal LOA, as they rely on an assumption of no association between delta and average of measurements (Bland et al., 1999; Ho, 2018). This also complicates the task of comparing these summary statistics with other papers in the literature, which have not reported their data in this detail. Future validation should consider reporting in such a way to compare data that contains biphasic magnitude-dependent bias more accurately, such as with regression-generated central tendency and between-case variability (Bland et al., 1999). Secondly, it remains uncertain what impact patients undergoing imaging at different imaging facilities 6 weeks after surgery may have had on the CT-derived data. The effect of multiple scanners on accuracy of CT-based landmark identification has not been reported for this specific application, however other CT-derived features have shown considerable variability between scanners (Varghese et al., 2019). In particular, variation in femoral rotation during image acquisition between the preoperative and postoperative scans may impact the ability to identify the longitudinal femoral axis, which is an important landmark for femoral offset. Therefore, it is plausible that some observer error could be attributed to the variation in scan quality. Future studies should consider quantifying this potential source of variation in clinical assessments.

## Conclusions

The navigation system assessed in a primary THA cohort for end-stage hip osteoarthritis provides acceptable validity within clinical recommendations for cup placement, femoral offset and leg length in the lateral decubitus patient position. This paper demonstrated that correction of magnitude-dependent biases observed for inclination and leg length change could further improve system accuracy in real-world application. The ability to apply accurate navigation systems to pathologic anatomy in the context of variable anthropometry remains an ongoing challenge for clinical practice.

## Data Availability

Upon reasonable request to the authors, only de-identified data (i.e. data with sensitive and personal information removed), will be provided as per the ANDS Publishing and Sharing Sensitive Data Guide.

## Acknowledgements

The authors acknowledge the participation of the patients and the contributions of the staff at Gold Coast Centre for Bone and Joint Surgery for the conduct of the study. The authors also acknowledge the contribution and dedication of Charles Day-Smith (South Coast Radiology) in the acquisition of imaging studies for the analysis. The authors also thank Meredith Harrison-Brown (EBM Analytics) for assistance with study ethics and assistance preparing the manuscript.

## Funding Declaration

This work was sponsored by Naviswiss AG. CS and MF declare a relationship through their employment (EBM Analytics) with the sponsor through a paid consultancy. TS declares a relationship through their employment (Medivation) with the sponsor through a paid consultancy. DL declares a relationship with the sponsor as an unpaid consultant to Naviswiss AG.

## Supplementary material

Supplementary material 1 - Code

Supplementary material 2 - Regression model results summary

Supplementary material 3 - Summary of validation findings

## Notes

### Competing Interest Statement

All authors have completed the ICMJE uniform disclosure form at www.icmje.org/coi_disclosure.pdf and declare the following:
CS and MF are employed by EBM Analytics, who were contracted by Naviwsiss AG to assist with completion of protocol manuscript and data analysis for study of same device (related to submitted work). In the past 36 months, EBM Analytics has received grants or contracts from OSSIS Ltd, Headsafe Ltd, DePuy Synthes Mitek Sports Medicine and Exactech Ltd for work unrelated to the submitted manuscript. CS is a shareholder in EBM Analytics.
DL received financial support from Naviswiss AG for completion of this study and support from Medivation for the image analysis component of this study. In the past 36 months, DL declares the following unrelated to the current work: grants or contracts from Zimmer Biomet and DePuy; royalties from Zimmer Biomet and DePuy; payment or honoraria for lectures, presentations, speakers bureaus, manuscript writing or educational events from Zimmer Biomet, DePuy and LifeHealthCare; support for attending meetings or travel from Zimmer Biomet, DePuy, Enovis; patent with DePuy, unpaid leadership roles with the Queensland AOA and the Asia Pacific Arthroplasty Society; stock in 360 MedCare and Naviwswiss AG; and an unpaid consultancy with Naviswiss AG.
TS's employer was contracted by Naviswiss AG to assist with the completion of the current study. TS additionally declares stock in Naviswiss AG.

### Clinical Trial

ACTRN12620000873921

### Funding Statement

This work was funded by Naviswiss AG. CS and MF declare a relationship through their employment (EBM Analytics) with the sponsor through a paid consultancy. TS declares a relationship through their employment (Medivation) with the sponsor through a paid consultancy. DL declares a relationship with the sponsor as an unpaid consultant to Naviswiss AG.

### Author Declarations

The Human Research Ethics Committee of Ramsay Health Care Queensland gave ethical approval for this work.

