## Supplementary material for "Imageless navigation system (Naviswiss) provides accurate component position in total hip arthroplasty with lateral decubitus position for end-stage hip osteoarthritis: A prospective cohort study with CT-validation": Supplementary material 1 - Code.pdf

```
**Tidy and load

clear

cd "..\Statistical Analysis"

use "NS Accuracy Analysis - Fourth Pass.dta"

import delimited "MasterAccuracy.csv"

** Summary statistics for patient characteristics
summarize ageatsurgery bmi delta_inc_fpp delta_ver_fpp delta_total_offset delta_total_lld ns_inc
ns_ver ns_offset ns_lld image_inc_fpp image_ver_fpp image_total_offset image_total_lld

*Fill missing BMI
summarize bmi

egen meanbmi = mean(bmi)

replace bmi = meanbmi if bmi ==.

****Part A - Calculate Agreement****

**bootstrap - calculate median of delta
bootstrap r(p50), reps(1000) seed(1234) nodots: summarize delta_inc_app, detail
bootstrap r(p50), reps(1000) seed(1234) nodots: summarize delta_inc_fpp, detail
bootstrap r(p50), reps(1000) seed(1234) nodots: summarize delta_ver_app, detail
bootstrap r(p50), reps(1000) seed(1234) nodots: summarize delta_ver_fpp, detail
bootstrap r(p50), reps(1000) seed(1234) nodots: summarize delta_total_offset, detail
bootstrap r(p50), reps(1000) seed(1234) nodots: summarize delta_total_lld, detail

**Rock out with the bootstrap out - calculate mean of delta
bootstrap r(mean), reps(1000) seed(1234) nodots: summarize delta_inc_app, detail
bootstrap r(mean), reps(1000) seed(1234) nodots: summarize delta_inc_fpp, detail
bootstrap r(mean), reps(1000) seed(1234) nodots: summarize delta_ver_app, detail
bootstrap r(mean), reps(1000) seed(1234) nodots: summarize delta_ver_fpp, detail
bootstrap r(mean), reps(1000) seed(1234) nodots: summarize delta_total_offset, detail
bootstrap r(mean), reps(1000) seed(1234) nodots: summarize delta_total_lld, detail

**Rerun with absolute deviations (all declarations included)****

generate abs_delta_inc_app = abs(delta_inc_app)
generate abs_delta_inc_fpp = abs(delta_inc_fpp)
generate abs_delta_ver_app = abs(delta_ver_app)
generate abs_delta_ver_fpp = abs(delta_ver_fpp)
generate abs_delta_offset = abs(delta_total_offset)
generate abs_delta_lld = abs(delta_total_lld)

bootstrap r(mean), reps(1000) seed(1234) nodots: summarize abs_delta_inc_app, detail
bootstrap r(mean), reps(1000) seed(1234) nodots: summarize abs_delta_inc_fpp, detail
bootstrap r(mean), reps(1000) seed(1234) nodots: summarize abs_delta_ver_app, detail
bootstrap r(mean), reps(1000) seed(1234) nodots: summarize abs_delta_ver_fpp, detail
bootstrap r(mean), reps(1000) seed(1234) nodots: summarize abs_delta_offset, detail
bootstrap r(mean), reps(1000) seed(1234) nodots: summarize abs_delta_lld, detail


**Are the differences significantly different to Zero?
signrank delta_inc_app = 0
signrank delta_inc_fpp = 0
signrank delta_ver_app = 0
signrank delta_ver_fpp = 0
signrank delta_total_offset = 0
signrank delta_total_lld = 0
```

```

**Work out if ave is associated with delta_inc_app

bootstrap, reps(100) seed(123): regress delta_inc_fpp ave_inc_fpp
bootstrap, reps(100) seed(123): regress delta_inc_app ave_inc_app
bootstrap, reps(100) seed(123): regress delta_ver_fpp ave_ver_fpp
bootstrap, reps(100) seed(123): regress delta_ver_app ave_ver_app
bootstrap, reps(100) seed(123): regress delta_total_offset ave_total_offset
bootstrap, reps(100) seed(123): regress delta_total_lld ave_total_lld

mkdir "\Statistical Analysis\FourthPass"
mkdir "\FourthPass\Original"
cd "FourthPass\Original"

*Plot what would be Bland_Altman plots
*95%CI is prediction interval for individual datapoint (not the mean)
* line coordinates specified are retrieved from LOA calculations listed in [AnalysisTables] in NS Dashboard sheet

twoway (lfitci delta_inc_fpp ave_inc_fpp, stdf acolor(gs6%50)) (scatter delta_inc_fpp ave_inc_fpp), yline(-8.0 10, lwidth(1pt) lcolor(blue) lpattern(dash))
graph export BlandAltmanInc.png, replace
graph close Graph
twoway (lfitci delta_ver_fpp ave_ver_fpp, stdf acolor(gs6%50)) (scatter delta_ver_fpp ave_ver_fpp), yline(-6.8 10.8, lwidth(1pt) lcolor(blue) lpattern(dash))
graph export BlandAltmanVer.png, replace
graph close Graph
twoway (lfitci delta_total_offset ave_total_offset, stdf acolor(gs6%50)) (scatter delta_total_offset ave_total_offset),yline(-4.0 7.7, lwidth(1pt) lcolor(blue) lpattern(dash))
graph export BlandAltmanOffset.png, replace
graph close Graph
twoway (lfitci delta_total_lld ave_total_lld, stdf acolor(gs6%50)) (scatter delta_total_lld ave_total_lld),yline(-5.2 5.6, lwidth(1pt) lcolor(blue) lpattern(dash))
graph export BlandAltmanLLD.png, replace
graph close Graph

cd "G:\My Drive\EBMA\Client Drive\Naviswiss\EBMA Working\Publications\DL Accuracy - Lateral Approach\Statistical Analysis"

save "NS_DL Accuracy Analysis - Fourth Pass.dta", replace

***Part B factors associated with agreement: *No declarations omitted

* encode categorical variables

rename gender sex
encode sex, gen(sexcode)


mkdir "\FourthPass\Original\Regression"
cd "\FourthPass\Original\Regression"

*Inclination
bootstrap, reps(100) seed(1234) nodots: regress delta_inc_fpp ns_inc c.bmi c.ageatsurgery i.sexcode, vce(robust)

margins sexcode

predict delta_inc_predict, xb


generate new_ns_inc = ns_inc - delta_inc_predict
generate delta_inc_biasc = new_ns_inc - image_inc_fpp
```

```
generate abs_delta_inc_biasc = abs(delta_inc_biasc)

bootstrap r(mean), reps(1000) seed(1234) nodots: summarize delta_inc_biasc, detail
bootstrap r(mean), reps(1000) seed(1234) nodots: summarize abs_delta_inc_biasc, detail

twoway (lfitci delta_inc_biasc ave_inc_fpp, stdf acolor(gs6%50)) (scatter delta_inc_biasc ave_inc_fpp),yline(-7.5 7.5, lwidth(1pt) lcolor(blue) lpattern(dash))
graph export BlandAltmanIncBC.png, replace
graph close Graph

*Version
bootstrap, reps(100) seed(1234) nodots: regress delta_ver_fpp ns_ver c.bmi c.ageatsurgery i.sexcode, vce(robust)

predict delta_ver_predict, xb

generate new_ns_ver = ns_ver - delta_ver_predict
generate delta_ver_biasc = new_ns_ver - image_ver_fpp

generate abs_delta_ver_biasc = abs(delta_ver_biasc)

bootstrap r(mean), reps(1000) seed(1234) nodots: summarize delta_ver_biasc, detail
bootstrap r(mean), reps(1000) seed(1234) nodots: summarize abs_delta_ver_biasc, detail

twoway (lfitci delta_ver_biasc ave_inc_fpp, stdf acolor(gs6%50)) (scatter delta_ver_biasc ave_inc_fpp),yline(-8.1 8.1, lwidth(1pt) lcolor(blue) lpattern(dash))
graph export BlandAltmanVerBC.png, replace
graph close Graph

*Offset
bootstrap, reps(100) seed(1234) dots: regress delta_total_offset ns_offset c.bmi c.ageatsurgery i.sexcode, vce(robust)

predict delta_offset_predict, xb

regress delta_offset_predict image_total_offset

generate new_ns_offset = ns_offset - delta_offset_predict
generate delta_offset_biasc = new_ns_offset- image_total_offset

generate abs_delta_offset_biasc = abs(delta_offset_biasc)

bootstrap r(mean), reps(1000) seed(1234) nodots: summarize delta_offset_biasc, detail
bootstrap r(mean), reps(1000) seed(1234) nodots: summarize abs_delta_offset_biasc, detail

twoway (lfitci delta_offset_biasc ave_total_offset, stdf acolor(gs6%50)) (scatter delta_offset_biasc ave_total_offset),yline(-4.9 4.9, lwidth(1pt) lcolor(blue) lpattern(dash))
graph export BlandAltmanOffsetBC.png, replace
graph close Graph

*LLD
bootstrap, reps(100) seed(1234) dots: regress delta_total_lld ns_lld c.bmi c.ageatsurgery i.sexcode, vce(robust)

predict delta_lld_predict, xb

generate new_ns_lld = ns_lld - delta_lld_predict
generate delta_lld_biasc = new_ns_lld - image_total_lld

generate abs_delta_lld_biasc = abs(delta_lld_biasc)

bootstrap r(mean), reps(1000) seed(1234) nodots: summarize delta_lld_biasc, detail
```

```
bootstrap r(mean), reps(1000) seed(1234) nodots: summarize abs_delta_lld_biasc, detail

twoway (lfitci delta_lld_biasc ave_total_lld, stdf acolor(gs6%50)) (scatter delta_lld_biasc
ave_total_lld),yline(-5.0 5.0, lwidth(1pt) lcolor(blue) lpattern(dash))
graph export BlandAltmanlldBC.png, replace
graph close Graph

** Summary statistics for absolute deviations
summarize abs_delta_inc_app abs_delta_inc_fpp abs_delta_ver_app abs_delta_ver_fpp abs_delta_off
abs_delta_lld

**Rerun with absolute deviations (all declarations included)****

**bootstrap - calculate mean of delta
bootstrap r(mean), reps(1000) seed(1234) nodots: summarize delta_inc_biasc, detail
bootstrap r(mean), reps(1000) seed(1234) nodots: summarize delta_ver_biasc, detail
bootstrap r(mean), reps(1000) seed(1234) nodots: summarize delta_offset_biasc, detail
bootstrap r(mean), reps(1000) seed(1234) nodots: summarize delta_lld_biasc, detail

bootstrap r(mean), reps(1000) seed(1234) nodots: summarize abs_delta_inc_biasc, detail
bootstrap r(mean), reps(1000) seed(1234) nodots: summarize abs_delta_ver_biasc, detail
bootstrap r(mean), reps(1000) seed(1234) nodots: summarize abs_delta_offset_biasc, detail
bootstrap r(mean), reps(1000) seed(1234) nodots: summarize abs_delta_lld_biasc, detail

*Declarations omitted
mkdir "FourthPass\DecOmit"
cd "\FourthPass\DecOmit"

* encode intraop declarations


generate str3 intraopdec2 = ""
replace intraopdec2 = "No" if intraopdec == "Nothing to declare"
replace intraopdec2 = "Yes" if !(intraopdec == "Nothing to declare")
encode intraopdec2, gen(deccode)

**Run analysis with declarations removed

generate delta_offset_decomit =.
generate delta_lld_decomit =.
generate ageatsurgery_decomit =.
generate sexcode_decomit =.
generate bmi_decomit =.
generate ave_offset_decomit =.
generate ave_lld_decomit =.
generate ns_offset_decomit =.
generate ns_lld_decomit =.

replace delta_offset_decomit = delta_total_offset if deccode ==1
replace delta_lld_decomit = delta_total_lld if deccode ==1
replace ageatsurgery_decomit = ageatsurgery if deccode ==1
replace sexcode_decomit = sexcode if deccode ==1
replace bmi_decomit = bmi if deccode ==1
replace ave_offset_decomit = ave_total_offset if deccode ==1
replace ave_lld_decomit = ave_total_lld if deccode ==1
replace ns_offset_decomit = ns_offset if deccode ==1
replace ns_lld_decomit = ns_lld if deccode ==1

**Rock out with the bootstrap out - calculate mean of delta
bootstrap r(mean) if !missing(delta_offset_decomit), reps(1000) seed(1234) nodots: summarize
delta_offset_decomit, detail
bootstrap r(mean) if !missing(delta_lld_decomit), reps(1000) seed(1234) nodots: summarize
delta_lld_decomit, detail
```

```
**Rerun with absolute deviations

generate abs_delta_offset_decomit = abs(delta_offset_decomit)
generate abs_delta_lld_decomit = abs(delta_lld_decomit)


*Plot Bland-Altman
twoway (lfitci delta_offset_decomit ave_offset_decomit, stdf acolor(gs6%50)) (scatter
delta_offset_decomit ave_offset_decomit),yline(-2.7 6.9, lwidth(1pt) lcolor(blue) lpattern(dash))
graph export BlandAltmanOffsetDecOmit.png, replace
graph close Graph
twoway (lfitci delta_lld_decomit ave_lld_decomit, stdf acolor(gs6%50)) (scatter delta_lld_decomit
ave_lld_decomit),yline(-4.3 5.0, lwidth(1pt) lcolor(blue) lpattern(dash))
graph export BlandAltmanLLDDecOmit.png, replace
graph close Graph

**bootstrap - calculate mean of delta
bootstrap r(mean), reps(1000) seed(1234) nodots: summarize delta_offset_decomit, detail
bootstrap r(mean), reps(1000) seed(1234) nodots: summarize delta_lld_decomit, detail

bootstrap r(mean), reps(1000) seed(1234) nodots: summarize abs_delta_offset_decomit, detail
bootstrap r(mean), reps(1000) seed(1234) nodots: summarize abs_delta_lld_decomit, detail

*12-Nov-2022


cd "G\Statistical Analysis"

save "NS Accuracy Lateral.dta", replace

export delimited "NS Accuracy Lateral - Output"

clear
```
