## Supplementary material for "Imageless navigation system (Naviswiss) provides accurate component position in total hip arthroplasty with lateral decubitus position for end-stage hip osteoarthritis: A prospective cohort study with CT-validation": Supplementary material 2 - Regression model summaries.pdf

Supplementary material 2 - Regression Models not used

Table 1: Regression summary delta inclination (FPP)

Linear regression

| | |
|---|---|
| Number of obs = | 33 |
| Replications = | 100 |
| Wald chi2(4) = | 6.72 |
| Prob > chi2 = | 0.1517 |
| R-squared = | 0.2092 |
| Adj R-squared = | 0.0963 |
| Root MSE = | 4.4274 |

| delta_in~fpp | Observed coefficient | Bootstrap std. err. | z | P>\|z\| | Normal-based [95% conf. | interval] |
|---|---|---|---|---|---|---|
| ns_inc | .2209985 | .5602995 | 0.39 | 0.693 | -.8771683 | 1.319165 |
| bmi | -.2109395 | .13427 | -1.57 | 0.116 | -.4741039 | .052225 |
| ageatsurgery | -.1249936 | .1127633 | -1.11 | 0.268 | -.3460056 | .0960185 |
| sexcode | | | | | | |
| M | -3.197874 | 1.621098 | -1.97 | 0.049 | -6.375169 | -.0205797 |
| _cons | 8.272596 | 27.49402 | 0.30 | 0.764 | -45.61469 | 62.15988 |

Table 2: Marginal Estimates - Delta Inclination and Sex

| | Margin | Delta-method std. err. | z | P>\|z\| | [95% conf. | interval] |
|---|---|---|---|---|---|---|
| sexcode | | | | | | |
| F | 2.535333 | 1.204659 | 2.10 | 0.035 | .174245 | 4.896421 |
| M | -.6625413 | .9833888 | -0.67 | 0.500 | -2.589948 | 1.264865 |

Table 3: Regression summary delta version (FPP)

Linear regression

| | |
|---|---|
| Number of obs | = 33 |
| Replications | = 100 |
| Wald chi2(4) | = 10.09 |
| Prob > chi2 | = 0.0389 |
| R-squared | = 0.1823 |
| Adj R-squared | = 0.0655 |
| Root MSE | = 4.2220 |

| delta_ve~fpp | Observed coefficient | Bootstrap std. err. | z | P>\|z\| | Normal-based [95% conf. | interval] |
|---|---|---|---|---|---|---|
| ns_ver | .4058231 | .1456059 | 2.79 | 0.005 | .1204408 | .6912054 |
| bmi | .0838532 | .1675331 | 0.50 | 0.617 | -.2445056 | .412212 |
| ageatsurgery | .042833 | .0876892 | 0.49 | 0.625 | -.1290347 | .2147007 |
| sexcode | | | | | | |
| M | -1.069394 | 1.513191 | -0.71 | 0.480 | -4.035193 | 1.896405 |
| _cons | -11.15331 | 8.028162 | -1.39 | 0.165 | -26.88822 | 4.581602 |

Table 4: Regression summary delta total offset

Linear regression

| | |
|---|---|
| Number of obs | = 33 |
| Replications | = 100 |
| Wald chi2(4) | = 8.13 |
| Prob > chi2 | = 0.0871 |
| R-squared | = 0.3269 |
| Adj R-squared | = 0.2308 |
| Root MSE | = 2.5558 |

| delta_tota~t | Observed coefficient | Bootstrap std. err. | z | P>\|z\| | Normal-based [95% conf. | interval] |
|---|---|---|---|---|---|---|
| ns_offset | .3873898 | .152568 | 2.54 | 0.011 | .0883621 | .6864175 |
| bmi | -.1056797 | .0965304 | -1.09 | 0.274 | -.2948758 | .0835164 |
| ageatsurgery | .0361236 | .0486822 | 0.74 | 0.458 | -.0592917 | .131539 |
| sexcode | | | | | | |
| M | 1.436563 | .9589039 | 1.50 | 0.134 | -.4428545 | 3.31598 |
| _cons | .9346356 | 3.761913 | 0.25 | 0.804 | -6.438579 | 8.30785 |

Table 5: Regression summary delta leg length difference

```
Linear regression                                Number of obs =         33
                                                 Replications  =        100
                                                 Wald chi2(4)  =       3.95
                                                 Prob > chi2   =     0.4121
                                                 R-squared     =     0.1417
                                                 Adj R-squared =     0.0190
                                                 Root MSE      =     2.7817
```

| delta_tota~d | Observed coefficient | Bootstrap std. err. | z | P>\|z\| | Normal-based [95% conf. | interval] |
|---|---|---|---|---|---|---|
| ns_lld | .3422847 | .2039923 | 1.68 | 0.093 | -.0575329 | .7421023 |
| bmi | -.1579938 | .1404499 | -1.12 | 0.261 | -.4332704 | .1172829 |
| ageatsurgery | -.0949432 | .0825915 | -1.15 | 0.250 | -.2568195 | .0669332 |
| sexcode | | | | | | |
| M | -.1577911 | .9626075 | -0.16 | 0.870 | -2.044467 | 1.728885 |
| _cons | 9.186773 | 8.249123 | 1.11 | 0.265 | -6.98121 | 25.35476 |
