## Supplementary material for "Imageless navigation system (Naviswiss) provides accurate component position in total hip arthroplasty with lateral decubitus position for end-stage hip osteoarthritis: A prospective cohort study with CT-validation": Supplementary material 3 - Summary of validation findings.pdf

| | This study | This study (absolute) | Hasegawa et al 2022[1] (supine - absolute) | Pooled[1-8] (N = 688) |
|---|---|---|---|---|
| Inclination_ FPP (°) | 1.0 (4.6) | 3.6 (3.1) | | |
| Inclination (Bias Corrected) | 0 (4.0) | 3.2 (2.7) | 2.8 (2.2) | 2.8 (2.0) |
| Version_ FPP (°) | 2.0 (4.5) | 4.0 (2.6) | | |
| Version (Bias Corrected) | 0 (4.0) | 3.4 (2.2) | 2.8 (2.0) | 3.6 (3.5) |
| Offset (mm)* | 2.1 (2.4) | 2.4 (2.1) | | |
| LLD (mm)* | 0.4 (2.4) | 1.8 (1.3) | | |

*Declarations omitted
